# Rapid Development of a *De Novo* Convalescent Plasma Program in Response to a Global Pandemic: A Large Southeastern U.S. Blood Center’s Experience

**DOI:** 10.1101/2020.10.23.20217901

**Authors:** Rita Reik, Richard R. Gammon, Nancy Carol, Judith Smith, Martin Grable, Susan Forbes, Yanyun Wu, Lance Reed, Michael Rogers, Alicia Prichard, Scott Paul, George Scholl

## Abstract

**Background:** With no vaccine or treatment for SARS-CoV-2 and its associated disease, COVID-19, convalescent plasma from recovered COVID-19 (CCP) patients offered a potential therapy. In March of 2020, the United States (US) Food and Drug Administration (FDA) authorized CCP under emergency Investigational New Drug (eIND) exemption and an IRB-approved Expanded Access Program (EAP) to treat severe COVID-19. Hospital demand grew rapidly in the Southeastern US, resulting in backlogs of CCP orders. We describe a large US blood center’s (BC) rapid implementation of a CCP program in response to community needs.

**Study Design and Methods:** From April 2 to May 17, 2020 CCP was collected by whole blood or apheresis. Initial manual approaches to donor intake, collection and distribution were rapidly replaced with automated processes. All CCP donors and products underwent FDA-required screening and testing.

**Results:** 619 CCP donors (299 females, 320 males) presented for CCP donation [161 (25.6%) whole blood, 466 (74.1%) plasmapheresis] resulting in 1219 CCP units. Production of CCP increased as processes were automated and streamlined, from a mean of 11 donors collected/day for the first month to a mean of 25 donors collected/day in the subsequent two weeks. Backlogged orders were cleared, and inventory began to accumulate 4 weeks after project initiation.

**Conclusion:** The BC was able to implement an effective *de novo* CCP collection program within 6 weeks in response to a community need in a global pandemic. Documentation of the experience may inform preparedness for future pandemics.

## Introduction

The novel coronavirus known as Severe Acute Respiratory Syndrome Coronavirus 2 (SARS-CoV-2) emerged in December 2019, as an unusual outbreak of pneumonia in Wuhan, the capital of Hubei Province, China. The associated disease was named Coronavirus Disease 2019 (COVID-19) by the World Health Organization (WHO). Following the first report to WHO on December 31, 2019, SARS-Co-V-2 began spreading rapidly internationally. On January 30, 2020 WHO declared the SARS-CoV-2/COVID-19 outbreak to be a public health emergency of international concern and on March 11, 2020 deemed it a global pandemic.^1^ On January 22, 2020 the first confirmed case of COVID-19 in the US was identified in Washington State.^2^ As of July 5, 2020 the case fatality rate for COVID-19 was 4.6%.^3,4^

COVID-19 symptoms range from mild to life-threatening requiring intensive care and mechanical ventilation with supplemental oxygen.^5^ This life-threatening aspect of the disease has resulted in over-burdened healthcare systems both globally and in the US.^6^ Treatment is largely supportive. Favorable experience with convalescent plasma in other viral infections including coronavirus (SARS-CoV), Ebola, Middle Eastern Respiratory Syndrome (MERS) and avian influenza led to interest in using CCP as a treatment for COVID-19. Small early studies from China suggested clinical improvement with the use of CCP in COVID-19.^7,8^ In light of this, on March 24, 2020 the US Food and Drug Administration (FDA) authorized the use of CCP for treatment of serious or immediately life-threatening COVID-19 through the following pathways: clinical trials, single patient emergency Investigational New Drug (eIND) or under an Expande d Access Protocol (EAP) through the National Expanded Access Treatment Protocol.^9,10^ We herein describe a large US BC’s experience in implementing a CCP program.

## Materials and Methods

On Jan. 5, 2020, as a result of BC hemovigilance monitoring, Senior Leadership at the BC was alerted to the potential significance of SARS-CoV-2 via an internal email communication referencing an article of interest about a virus outbreak in Wuhan, China affecting 44 people.^11^ By early March, the pandemic started to significantly impact the BC’s service area. Shelter-in-place orders and social distancing measures began to affect blood collections, raising concerns about the adequacy of the regional blood supply. However, following the cancellation of elective surgeries at area hospitals in order to make beds available for the large influx of COVID-19 patients, the demand for traditional blood products dropped approximately 50%. On March 24, 2020, the BC launched a CCP implementation project with goals of meeting the need for CCP and building a robust inventory of frozen and liquid CCP to be made available for rapid fulfillment of orders. Recognizing the uniqueness and urgency of the situation, BC Senior Leadership committed to hands-on running of the project throughout the startup and additionally engaged an expert consultant who had experience with the COVID-19 outbreak in China to assist in the CCP program design and development.^12^ The BC Business Continuity Plan (BCP) had been previously activated in the context of the global pandemic. The Project Management Office (PMO) was enlisted to support design and management of the CCP project. Institutional Review Board approval was obtained (Advarra – Pro00038077) and existing BC staff were trained and redeployed into various aspects of the project.

The urgent clinical need for CCP necessitated a two-phased approach to implementation of CCP production. Phase 1 (March 24 to April 27, 2020) involved primarily manual processes. During Phase 2 (April 28 to May 17, 2020) progressive process streamlining and automation enhanced CCP donor recruitment, intake, scheduling, and distribution. (Figure 1)

**Figure 1.**
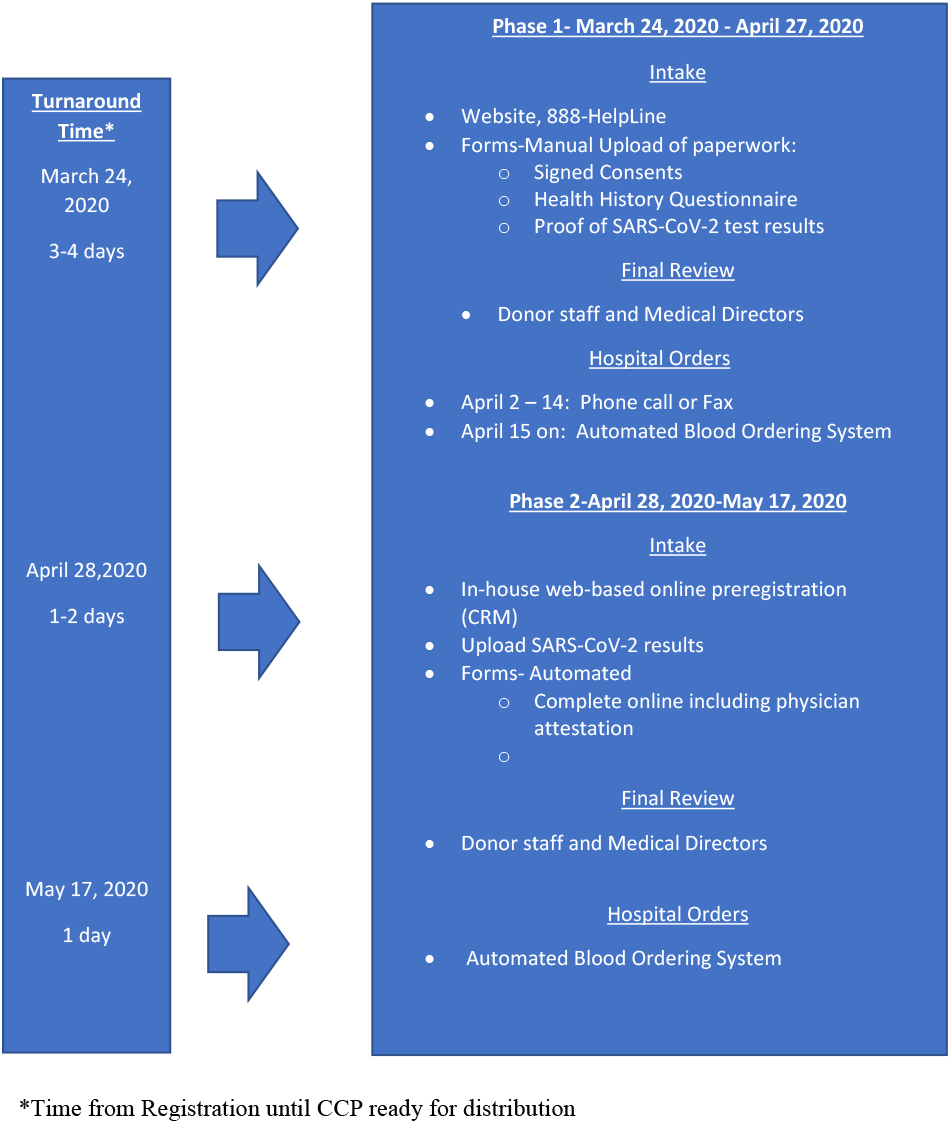
Overview of CCP Program Development

### Phase 1 (Manual Process) March 24 – April 27, 2020

#### Recruitment

On March 27, 2020, the BC informed contracted hospitals of its intention to produce CCP and asked for their assistance in identifying potential CCP donors. A recruitment website (Figures 2,3) and encrypted email were developed and by March 30, 2020 an 888-HelpLine was launched for intake and manual tracking of potential CCP donors. A coordinated media and public relations campaign were enacted in conjunction with an integrated social media appeal to create awareness for the urgent need for people who recovered from COVID-19 to register with the BC to be a CCP donor. The Florida Department of Health (FDOH), the Florida Governor’s office and the Florida Medical Association (FMA) assisted in identifying CCP donors. On April 2, 2020, the first unit of CCP was collected from whole blood and transfused as liquid plasma the next day.

**Figure 2.**
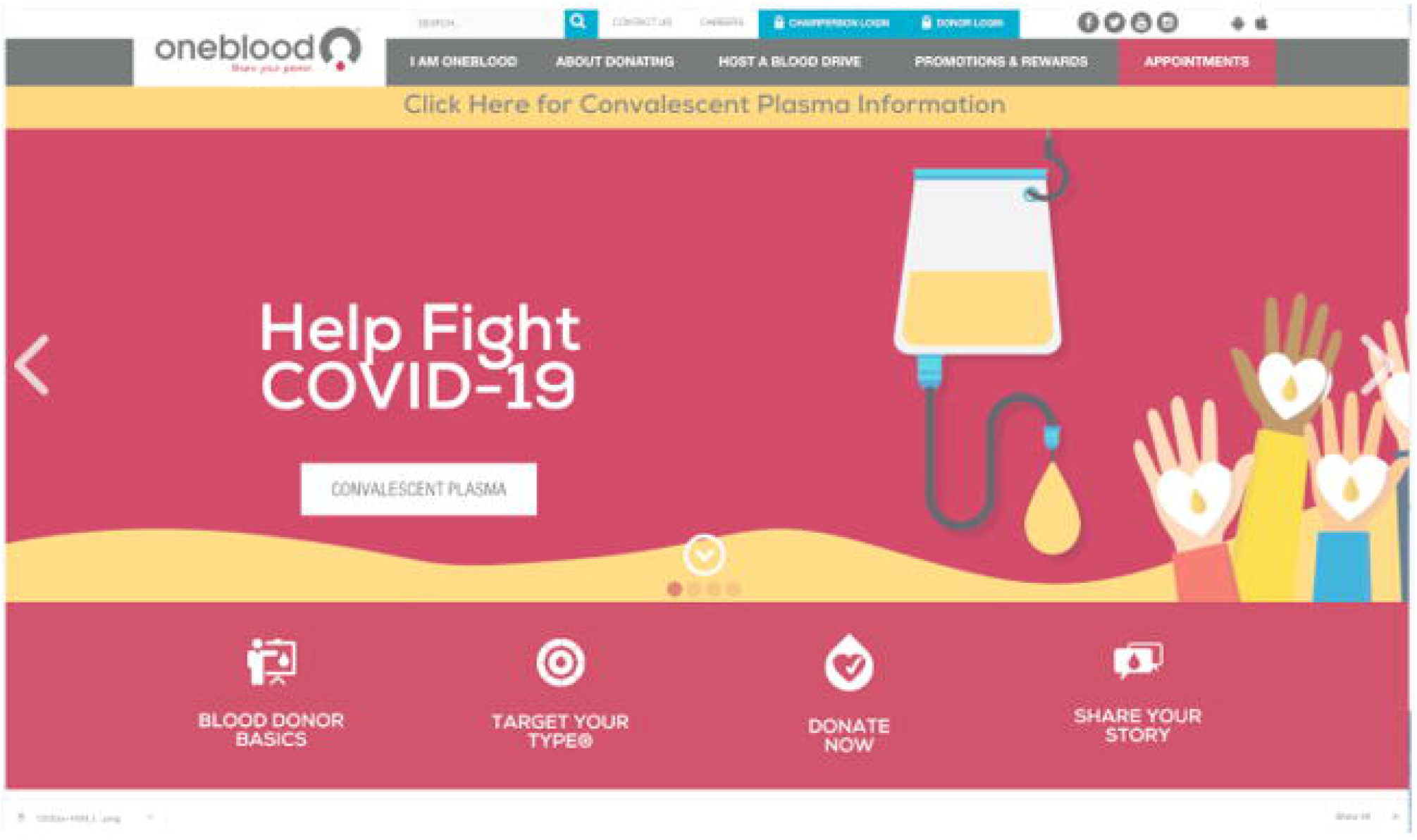
BC CCP Website Landing Page

**Figure 3.**
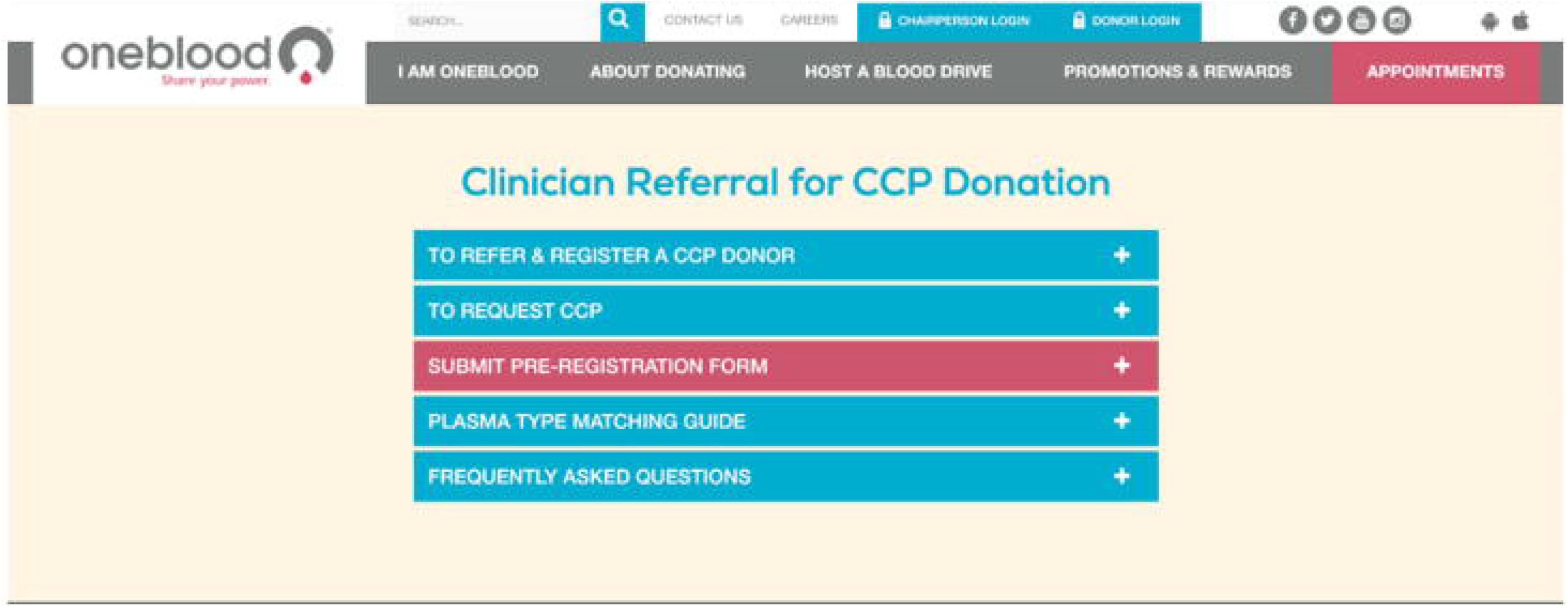
BC CCP Website Physician Landing Page

Donors presenting during Phase 1 consisted primarily of recovered COVID-19 patients identified by physicians or family members, who volunteered as directed donors for those with life-threatening COVID-19. Eligible patients had to be identified to the BC by their physician by means of an eIND or EAP patient number. During the first week of the project, most hospitals and physicians ordered CCP using the eIND pathway and obtained approval directly through the FDA’s Office of Emergency Operations.^10^ BC staff were available continuously throughout this period to field questions from donors and physicians, receive orders from hospitals, and communicate with FDA, FDOH and FMA. CCP donors were recruited according to FDA criteria (Table 1).^9^ Potential donors and/or their referring physicians were directed to the BC CCP recruitment website, 888-HelpLine or encrypted email where they completed consents, a health history questionnaire form and uploaded their SARS-CoV-2 laboratory reports. Donor intake information was reviewed by BC staff and forwarded to the BC Medical Directors for approval. Copies of laboratory testing were verified, and laboratory test sites and tests used were checked for acceptability using the FDA website as a reference.^13^ Following successful intake, pre-screening of the full donor history questionnaire was administered by telephone. The prescreen was intended to optimize blood center collection capacity and staffing, and compliance with “shelter-in-place” orders by increasing the probability that those donors who presented would be qualified for CCP donation.

**Table 1.**
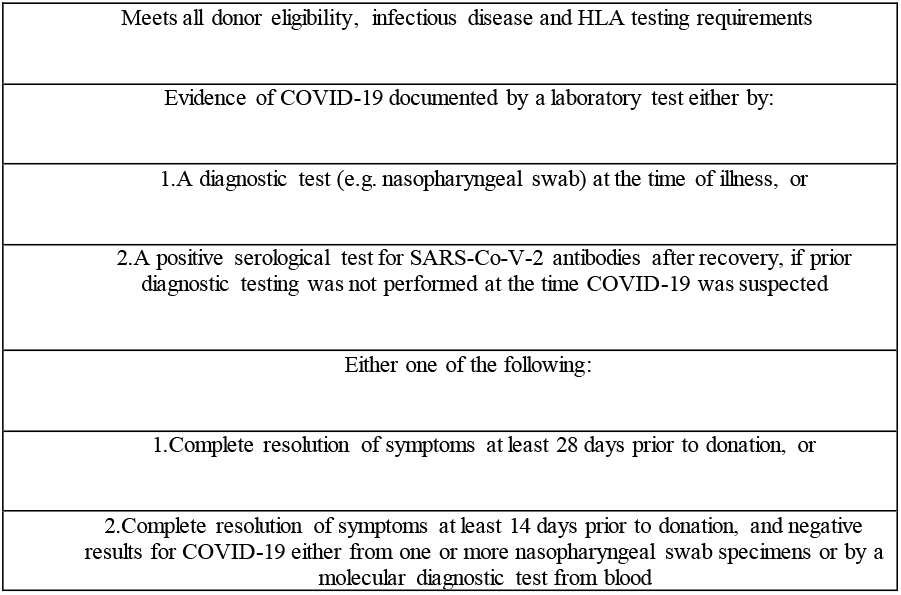
US FDA’s CCP Donor Criteria

#### Collection

Donors passing initial screening steps were scheduled for the earliest available appointment. Collections took place on six dedicated mobiles in a setting that observed all social distancing precautions and personal protective equipment (PPE) recommendations made by the Centers for Disease Control and Prevention (CDC) regarding masks, gloves, surface disinfection, and a minimum of six feet (2 meters) of distancing between persons. Each mobile was designed to accommodate twelve donors per day and only one donor was collected during one appointment time. Products were collected as whole blood (all first-time donors) or plasmapheresis (repeat donors) depending on donor preference and staff assessment.

Whole blood plasma collection was performed using Haemonetics bags (Haemonetics, Braintree, Mass.). Apheresis plasma collection was performed using Fresenius Kabi Alyx equipment with Fresenius Kabi 4R5730 bags (Fresenius Kabi USA, Lake Zurich, IL), and Terumo BCT TRIMA equipment with TRIMA set plasma/RBC kit, type 80500 (Terumo BCT, Lakewood, CO) instrumentation following the manufacturer’s recommendations. At the time of collection, units were tagged as CCP for immediate processing upon arrival at the BC’s manufacturing laboratories.

#### Laboratory Testing

All CCP donors underwent US FDA required transfusion-transmitted infectious disease (RTTID) and red cell antibody testing. Females with a history of pregnancy were tested for human leukocyte antigen (HLA) antibodies. Units testing positive for any of the required tests were not used for transfusion.

#### Processing, Labeling and Distribution

Storage and manufacturing of plasma from whole blood and apheresis was performed following the BC’s standard operating procedures. The resulting plasma products were labeled and tagged according to FDA requirements and shipped as liquid or frozen depending on the urgency of the situation. Products were manually tracked and delivered to hospitals as soon as required testing was completed and confirmed acceptable.

#### Ordering and Invoicing

From April 2-14, 2020, orders were placed by phone or fax. From April 15, 2020 forward, orders were placed through the BC automated blood ordering system software (BloodHub, Phoenix, AZ)). Cost recovery for units distributed under the EAP was reimbursed by the Biomedical Advanced Research Development Authority (BARDA) and were issued free of charge to hospitals. Units issued under eIND were charged to hospitals using the cost recovery model.

#### Quality Assurance

Rapidly changing information from regulatory and accrediting agencies made it imperative that the BCP team implemented the most current requirements in a controlled and compliant manner. This was accomplished by directing senior Quality Assurance (QA) staff and Medical Directors to audit and oversee the new processes. Labeling, licensing, interstate shipment, look-back and notification information was implemented in accordance with FDA requirements, however the fluidity of the situation did not allow time for necessary Blood Establishment Computer System changes and controls, resulting in the need for manual workarounds and planned deviations involving standard operating procedures, other documents and related training.

#### Customer Relations and Communication

All communications were coordinated with the marketing team to ensure consistent messaging. Concerted attempts were made to assure ongoing and proactive CCP inventory communications with hospitals and physicians. In addition to regular email communications with hospitals, the BC Chief Medical Officer (CMO), Executive Vice President and Medical Director gave multiple SARS-CoV-2/COVID-19 updates to the FMA Board of Directors. In addition, the CMO initiated a series of communications with the American Hospital Association (AHA), the American Medical Association (AMA) and America’s Blood Centers (ABC) regarding the need for CCP donors. The Chief Executive Officer (CEO) and CMO were also in close contact with the Florida Governor’s office and the State Surgeon General, providing frequent updates focused on the critical need for CCP donors, the status of CCP inventory and COVID-19 epidemiologic research.

#### Employee Relations and Communications

In late-March 2020, in anticipation of changes in working conditions associated with the pandemic, orders were placed for clean room coveralls, shoe coverings, lab coats, surgical masks, protective goggles and red bio-disposal bags for front line collections and therapeutic apheresis team members. Orders for gloves and cleaning supplies were increased. No staff were furloughed, but staff hours were flexed, and staff redeployed as needed to accommodate the rapidly changing needs. All administrative staff were directed to work remotely and Mission (Hazard) Pay incentive was implemented for hourly employees and increased on March 29, 2020. Employees were informed via frequent email updates and the company Intranet which hosted two informationa l pages; one with all employee related updates and frequently asked questions and the second with educational information and communications regarding the BC’s CCP processes. Daily meetings of the BCP and ad hoc project teams were also conducted.

#### Media Relations and Communications

On April 2, 2020, the CCP communications plan was implemented when the first CCP donor, Miami Mayor donated his plasma. The next morning, Communications issued a press release to media outlets that included an interview with Mr. Suarez encouraging others who had recovered from COVID-19 to donate their plasma. The story was also posted on BC social media channels. The BC Communications team gathered additional stories with more than a dozen of the initial CCP donors and continued to post on social and traditional news media. In addition, the BC created videos that took viewers behind the scenes to show the efforts taking place to collect, test and distribute CCP. A BC public service announcement (PSA) aired on local television stations and cable outlets that brought additional awareness to the need for more CCP donors. All materials were also displayed on the BC website and CCP donor registration page. Communications with FDA, other BCs, and national associations (e.g. ABC, AABB) and organizations (e.g. Blood Centers of America) reached a high level of intensity as information was exchanged regarding the implementation of CCP.

#### Phase 2 (Automated Process) April 28, 2020 – May 17, 2020

Conversion of the intake process from manual to automated involved using an internally developed web-based donor pre-registration solution and Microsoft’s (Microsoft, Redmond, WA) Customer Relationship Management (CRM) Software. The internally developed solution allowed for CCP donor pre-registration through a semi-automated Health History Questionnaire (HHQ) that the donor could download, populate and upload along with the required test results and Health Insurance Portability and Accountability Act (HIPAA) acknowledgement. The HHQ captured donor demographic information in addition to specific SARS-CoV-2 test dates, results and the donor’s blood type if known. Donors and clinicians could direct a donation to a specific patient by providing a de-identified patient eIND or EAP number. The CRM system automatically received the web entered pre-registration information for each donor and tracked CCP donors through review, Medical Director approval and donation scheduling.

In addition, by April 15, 2020, the inventory/distribution aspect was automated via blood ordering system software that accepted orders for CCP from the BC’s hospital partners. Each order required a patient eIND or EAP number and blood type. CRM automated reporting of the collections throughout the day provided advance notice of incoming components. This allowed for efficient planning of product allocation. It also allowed for real-time updates to the hospitals and clinicians caring for COVID-19 patients. Streamlining the intake process resulted in elimination of the HHQ on April 28, 2020, in favor of a simpler, fully automated form that could be completed online designed using Click Dimensions software (Click Dimensions, Atlanta, GA). The new form included a requirement to provide the date of the last COVID-19 symptom. Verification of FDA Emergency Use Authorization (EUA)-approved diagnostic testing by means of uploaded laboratory test record review was performed, however as an alternative, physician attestation of the donor’s required testing and results could be completed online. In early April 2020, the turnaround time from donor web pre-registration to scheduled donation spanned several days. With the automated process implemented on April 28, 2020 turnaround time was reduced to as low as one day in cases where the donor submitted all information correctly and was available via phone to schedule their donation appointment when first contacted.

## Results

From Apri1 2, 2020 through May 17, 2020 there were 619 unique CCP donors and 1219 CCP products produced with the characteristics described in Table 2. There was progressive loss of donors and product throughout the process (Figure 4). Unique donation discards due to specific test results are listed in Table 3. In comparison to the manual Phase 1, CCP production and distribution increased significantly during automated Phase 2 of the project, ranging from means of 11 donors collected and 18 products distributed per day in Phase 1 to 25 donors collected and 25 units distributed per day in Phase 2. The backlog of patient orders was cleared, and inventory was able to be built less than 4 weeks after initiation of the project (Figures 5,6 and Table 4).

**Table 2.** CCP Donor and Product Characteristics

| **For records dated April 2 through May 17, 2020:** | **Values** |
| --- | --- |
| Count of males | 320 |
| Count of females | 299 |
| Mean age Males | 48 |
| Mean age Females | 44 |
| First time OneBlood donors | 432 |
| Repeat OneBlood donors | 187 |
| Number of first-time CCP donors | 619 |
| Number of repeat CCP donors | 8 |
| Number and percentage of WB | 161 (25.6%) |
| Number and percentage of Plasma only | 466 (74.1%) |
| Mean mL of plasma volume collected from WB | 342 |
| Mean mL of plasma volume collected from plasma only collections | 649 |
| Total Products | 1219 |
| Number of products discarded due test to results | 111 |
| Number and percentage of products discarded due to breakage | 9 |
| Number and percentage of products discarded due to all other reasons | 17 |
| Total (%) products discarded | 137 (12%) |

**Table 3.** CCP donations discarded due to test results

| **Test** | **Number Positive** |
| --- | --- |
| Human Immunodeficiency Virus 1 and 2 Antibody (anti-HIV-1/2) | 1 |
| T. cruzi (Chagas) Antibody | 2 |
| Hepatitis B Core Antibody (anti-HBc) | 12 |
| Hepatitis C Virus Antibody | 5 |
| Human T-Cell Lymphotropic Virus I/II Antibody (anti-HTLV-I/II) | 1 |
| Serologic Test for Syphilis (STS) | 10 |
| Red Blood Cell Antibodies | 3 |
| HLA Antibodies | 77 |
| Total Discards Due to Testing | 111 |

**Table 4.**
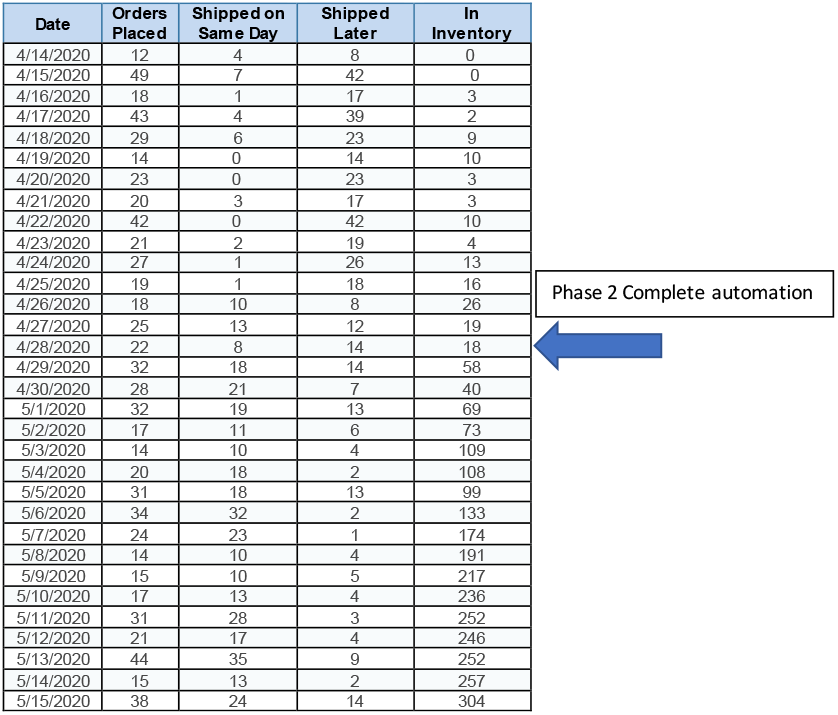
Phase 1 and Phase 2: Order fulfillment, turnaround time and inventory

**Figure 4.**
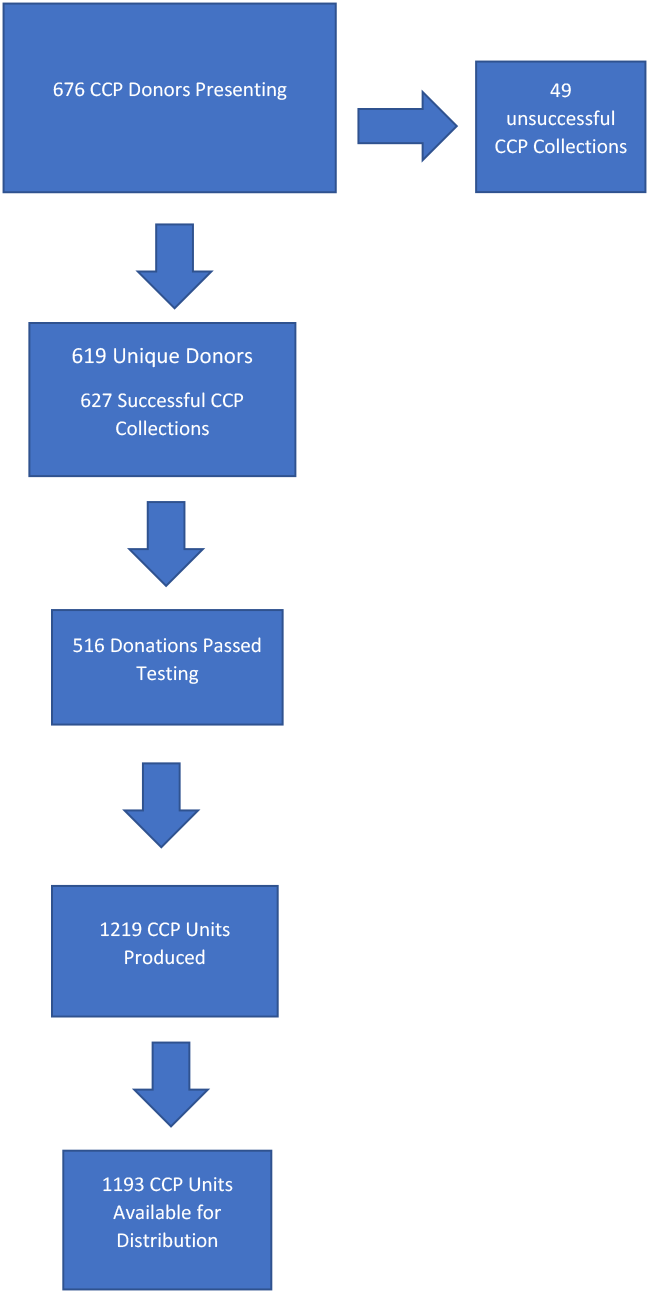
Progressive donor and product losses

**Figure 5.**
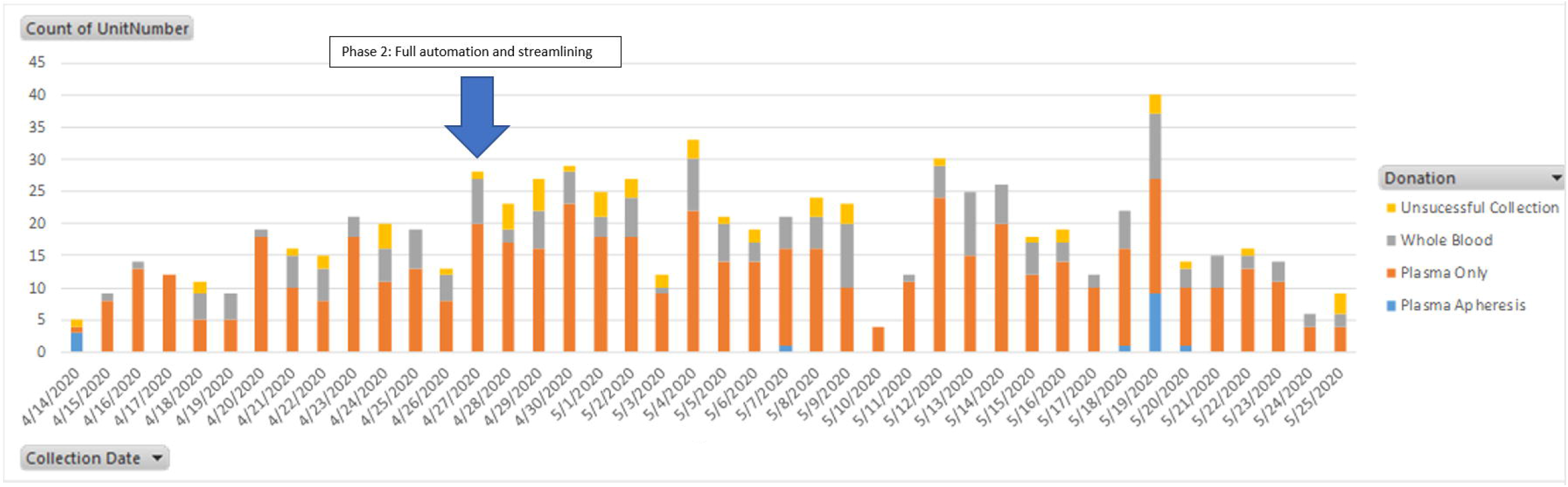
Breakout of CCP product type by date collected

**Figure 6.**
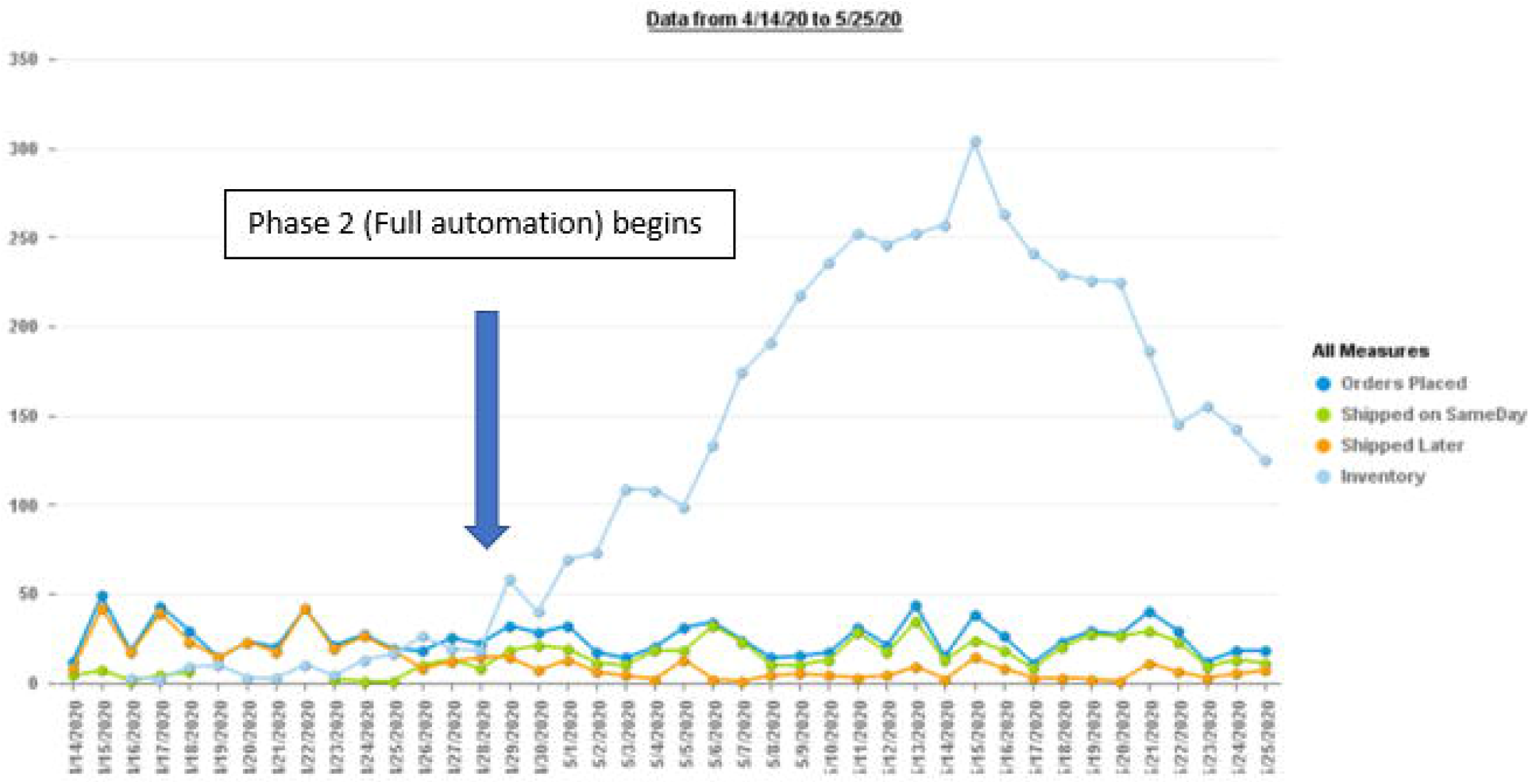
Phase 1 and Phase 2: Orders vs Shipments and Inventory by Date

## Discussion

We herein described the six-week implementation of a *de novo* CCP program which was able to identify and recruit qualified donors and track and timely distribute CCP units to patients with critical and life-threatening COVID-19. The CCP initiative described here had two aims: 1) to rapidly increase CCP production capacity to fulfill critical need; and 2) to build an inventory of CCP products in order to improve product delivery turnaround times. Based on the results for the timeframe described, we considered our goals to have been met. In reflecting on lessons learned throughout the process, the following key areas were identified:

### 1. Data

Comprehensive data was lacking regarding the regional and national blood supply, the number of units available in hospitals, and hospital usage patterns. This impaired the ability of the BC to predict the effect of the pandemic on the blood supply. As it impacted a large portion of the nation’s BCs simultaneously, it affected the BCs ability to move blood from unimpacted to impacted areas. Additionally, lack of data impaired the BCs ability to predict the effect of cancellation of elective procedures or the regional need for blood products, a capability which would have proven invaluable for integrating CCP production planning into operations.

### 2. Communication and Coordination

There were issues with communication at all levels, particularly nationally, highlighting the need for enhancing existing networks and communications pathways and keeping them open. Various parts of the nation were impacted in different ways and times by the pandemic, resulting in disjointed communications between regions and national organizations regarding prioritization of need for resources. The FDA and other national and regional organizations compensated for these gaps by making themselves available at all times. At the regional level, the BC Communication team’s proactive approach proved effective in controlling messaging to employees and the public.

### 3. Donor recruitment

CCP donor recruitment challenges were mainly attributable to the BC lack of access to qualified CCP donor lists. This resulted from regulations intended to protect patient privacy but had the unintended consequence of inhibiting the hospitals and health departments from sharing needed information about individuals who had recovered. This resulted in weeks of delay in CCP inventory development. Another donor recruitment lesson learned was related to timing. Initially, FDA required a 28-day deferral period after a CCP patient became asymptomatic. Since the epidemic was in its early stages in the US, many COVID-19 convalescent individuals had only recently recovered from their illness. Therefore, the 28 day wait period significantly limited the size of the eligible donor base. Another ongoing donor-related challenge was inconsistent national messaging regarding disease risk and mitigation.

### 4. Testing

The lack of licensed tests for SARS-CoV-2 presented challenges to donor recruitment and CCP product characterization. There was a plethora of new serologic and molecular tests on the market for SARS-CoV-2 that had received EUA by FDA, however lack of test availability remained an issue throughout the two phases. Trained staff were required staff to check the FDA website of EUA approved tests to determine if the test qualified.^13^ This process was an impediment to rapid donor intake and processing. Since there were no widely available antibody titer tests to verify CCP product efficacy the FDA recommended additional donor samples were to be collected and stored for future titer testing. The availability of extra sample tubes proved critical for later titer testing and research.

### 5. Personal Protective Equipment (PPE) and other supplies

The need for additional PPE put a strain on the BC at a time when there were national shortages. Severe supply chain shortages of PPE, disinfectants, paper goods and apheresis kits persisted throughout the project. This created pressure on BC administration to find needed supplies, an effort that was hampered by lack of prior relationships with vendors. Single source suppliers and just-in-time inventory practices, initially designed to reduce BC expenditures, exacerbated the situation.

### 6. Messaging

Inconsistent national messaging regarding risk, safety of blood donation and need for PPE resulted in ongoing confusion of donors and staff as to what social distancing measures were appropriate and resulted in the loss of significant numbers of donors and drive cancellations. Proactive communication strategies using social media, emails and traditional media helped to maintain open lines of dialogue with employees and the public, who were reassured by consistent messaging that the BC followed all recommended FDA and CDC guidelines for protective measures and that it was safe to donate.

### 7. Hospitals and Clinicians

Clinicians were unfamiliar with CCP resulting in hospitals and physicians being inadequately prepared for the rollout of the product. The unproven safety and effectiveness of CCP coupled with a lack of other therapeutic modalities for COVID-19 resulted in a level of uncertainty around availability and use of the product. This required ongoing communication between clinicians and BC physicians. Convalescent plasma had been used in previous viral outbreaks for decades, yet only in March of 2020 did FDA authorize it for BCs to collect and physicians to use under an eIND or EAP for treatment of COVID-19. The EAP and eIND pathways were unfamiliar to some non-research-based hospitals and BCs, which found the requirements cumbersome. The lack of coordination and preparation at the BC/hospital level with the national programs (eIND and EAP) and the need for use of manual systems during the early development of the program caused delays and frustration. Physician and public lack of education regarding the challenges faced in implementing a CCP program led to public relations issues for the BC, which was expected to be able to produce CCP on demand, like other blood products.

Another lesson learned was that while a CCP donor may present, there is no guarantee that a successful collection will occur. Even if CCP is obtained, ABO incompatibility might make the unit unsuitable. This was also a point of education for the ordering physicians who recruited family members to provide directed donations. Situations occurred in which family members were assured that the unit would be obtainable shortly after the donation only to discover later that the unit was unusable for various quality reasons.

### 8. BC Supporting Infrastructure

The pre-existence of a strong implementation support structure including the PMO, in-house IT/Business Intelligence (BI) and BCP teams prior to the onset of the pandemic greatly enhanced BC responsiveness to the CCP project challenges and proved to be critical in disaster management. The PMO at the BC consisted of a dedicated, experienced team of professionals who were rapidly deployed on the project to develop and revise processes and training in response to frequent changes in FDA requirements. This department played a pivotal role in ensuring the success of the project.

IT/BI was instrumental in streamlining and automating the intake and distribution aspects of the process. The BI team tracked and transformed complex data into highly functional dashboards and reports that allowed real time assessment and strategy development.

The BCP team performed daily horizon scanning on a global level and kept BC Leadership apprised of the progression of the pandemic and any additional threats. They gathered the CCP implementation team together daily for updates to facilitate and maintain communication in an extremely fluid environment.

## Conclusion

The SARS-CoV-2 global pandemic has unexpectedly thrust the blood banking community into the international spotlight and challenged it in a manner that was not fully anticipated, causing blood bankers to become acutely aware of the strengths and weaknesses in the system. The public and the government now look to the blood banking community for recommendations to ensure a safe and available blood supply at times when it is needed most urgently. This includes building on the strengths and correcting the gaps identified in sustaining the blood supply during this global pandemic.

Based on this BC experiences from March 24 through May 17, 2020 the following was concluded:

- There is an urgent need for reliable, real-time data on national blood inventory and usage granular to the regional level.
- The single supplier, just-in-time inventory model is not suitable for disasters. BCs would benefit from a stockpile of several weeks’ worth of critical supplies (e.g., PPE and collection kits), and relationships with multiple vendors in diverse locations.
- Established federal and state mechanisms are needed to allow public health departments, hospitals, and physicians the discretion to release select elements of protected patient information in the event of a significant disaster.
- The federal government should have mechanisms in place for streamlining processes for donor recruitment, testing and new product development in the event of another pandemic (or disaster) of significance to the nation’s blood supply. This would allow BCs to have emergency SOPs, IT pathways and training in place when needed.
- It is likely that there will be future outbreaks of viruses that may require treatment with CCP. Therefore, there should be standing federal protocols in place to allow rapid deployment of this product.
- In the event of a pandemic caused by an agent that is transfusion-transmissible, pathogen reduction of the entire blood supply would be required, therefore rapid development and consideration of funding of this capability should be a top priority.
- BCs benefit from robust, pre-existing, dedicated PMO, IT/BI and BCP capabilities in the event of a disaster.

The SARS-CoV-2 pandemic represents a unique event that has highlighted the need for planning for the unplannable. BCs recording their collective experiences with managing this pandemic will serve to better inform strategies for future disasters.

## Data Availability

There is no data available outside of this submitted manuscript.

## Acknowledgements

T Foster, B Mair, M Saint Martin, N Losa, K Risvold, E Rojas, M Lopez, C Shea, J Holder, M Bhuiyan, S Varghese all for their technical help with this manuscript.

